# Use Crow-AMSAA Method to predict the cases of the Coronavirus 19 in Michigan and U.S.A

**DOI:** 10.1101/2020.04.03.20052845

**Authors:** Yanshuo Wang

**Affiliations:** Reliability and Data Mining Consultant

## Abstract

The Crow-AMSAA method is used in engineering reliability world to predict the failures and evaluate the reliability growth. The author intents to use this model in the prediction of the Coronavirus 19 (COVID19) cases by using the daily reported data from Michigan, New York City, U.S.A and other countries. The piece wise Crow-AMSAA (CA) model fits the data very well for the infected cases and deaths at different phases while the COVID19 outbreak starting. The slope β of the Crow-AMSAA line indicates the speed of the transmission or death rate. The traditional epidemiological model is based on the exponential distribution, but the Crow-AMSAA is the Non Homogeneous Poisson Process (NHPP) which can be used to modeling the complex problem like COVID19, especially when the various mitigation strategies such as social distance, isolation and locking down were implemented by the government at different places.

**Summary:** This paper is to use piece wise Crow-AMSAA method to fit the COVID19 confirmed cases in Michigan, New York City, U.S.A and other countries.

## 1. Introduction

The COVID 19 was first found in Wuhan, Hubei Province, China in December, 2019, and it has been presented a main threat to the public health systems around the globe. As of April 12,2020, there have been about 1.8 million confirmed case, and about 116,000 reported deaths globally[3]. In U.S.A, there are about 561,159 confirm cases, and about 22,133 reported deaths [3]. In the state of Michigan, there are about 24,638 confirmed cases and about 1,487 reported death at the time author writing this paper [3][4]. The COVID19 is affecting 210 countries and territories around the world and 2 international conveyances. The COVID19 is spreading into all the 50 states, District of Columbia and its territories in United States. Because of the contagious of this disease, most of the states such as Michigan have issued the staying home order to reduce the infectious speed. The author has observed the U.S. and Michigan infected cases and deaths since March 16th. The author was curious that there must be a statistical model to predict this event. Since the Crow-AMSAA model is used for automotive warranty data by author to predict the failures numbers in the field. When the COVID19 infected data and death data were plugged in this model for Michigan, it is very surprised that it fits this model very well when the outbreak starts. Then the author continues to update this fitting by using the daily reported data from Michigan, and attempts to predict the next few day’s infected cases and deaths. On 3/28/2020, the author decided to plug all the U.S.A infected and death data in this model, it is also surprised that fits the Crow-AMSAA model as well. Then the author decided to write this paper to describe what the Crow-AMSAA model is and how the analysis has been done. The Crow-AMSAA model might be useful to predict the infected cases and deaths for a pandemic like COVID19. The daily reported data from New York City[12], Spain, Italy, France, Germany, UK, China and South Korea [3] have also been analyzed by using the piece wise Crow-AMSAA model. The comparison of the speed of the transmission and death rates at different places and countries are also summarized in this paper.

## 2. Review of Epidemiological Model

There are existing epidemiological models which used in the pandemic prediction.

### Exponential Model

It is believed that most epidemics grow approximately exponentially during the initial phase of an epidemic. *I(t)* is the number of diagnosis infected case, *t* is the time which is measured in days[5].

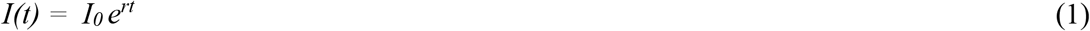

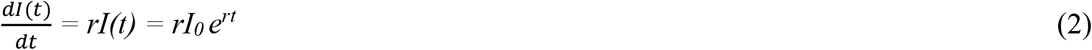

Where *r* is the growth rate, *I*_*0*_ is the constant which can be calculated by fitting the data.

### Susceptible-Infectious-Recovered (SIR) model

SIR model is the compartmental models which are used to simplify the mathematical modelling of infectious disease.

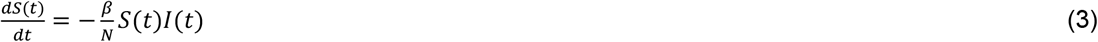

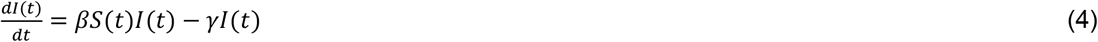

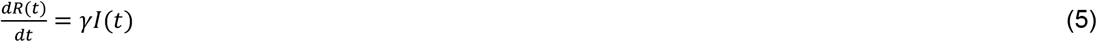

where *S(t)* is the number of susceptible individuals, *I*(t)is the number of infectious individuals, and *R(t)* is the number of recovered individuals; *β* is the transmission rate per infectious individual, and *γ* is the recovery rate, N is the population, *N = S(t)+I(t)+R(t)* [8].

### Logistic Model

Logistic model was developed by Belgian mathematician Pierre Verhulst (1838). Logistic model is the model which shows initially exponential growth followed a gradual slowing down and a saturation [8].

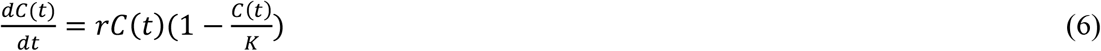

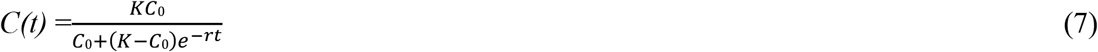

Where *C(t)* is the cumulative total numbers of infectious, *r* is the exponential growth rate, *K* is the upper limit of population growth and it is called carrying capacity. *C*_*0*_ is the *C(t)* when *t=0*

## 3. Crow-AMSAA Model

James T. Duane at GE Motors Division conducted the reliability growth analysis by observing the cumulative failure rates of the product subsystems during the development test. He plotted the cumulative failures versus the development time on a log-log paper (Duane, 1964). The AMSAA model, a major improvement in Duane’s approach was developed by Dr. Larry Crow in 1974 while he was at the Army Material Systems Analysis Activity (AMSAA). Dr. Crow proposed that the Duane model can be represented as non-homogeneous Poisson process (NHPP) model under Weibull intensity function [1][2].

The total confirmed infected case or deaths *N(t)* can be expressed as following when Crow-AMSAA model applies

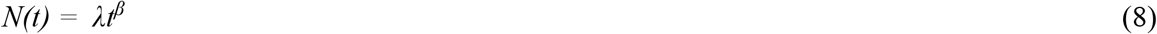

Where *t* is the time which measured in days, *λ* and *β* are constants, they will be explained later. The logarithm of cumulative events *N(t)* versus logarithm time *t*, which measured in days is a linear plot. By taking the natural logarithms of equation (8)

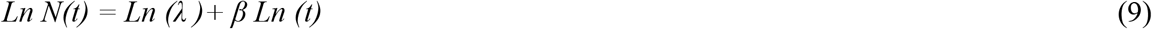

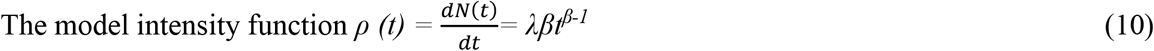

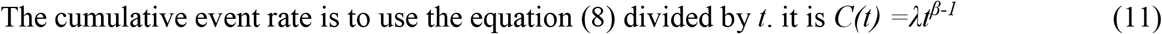

The intensity function is the derivative of the cumulative events *N(t) = λt*^*β*^, *ρ (t)* is called the rate of occurrence (ROC). In equation (9), the scale parameter, *λ*, is the intercept on the *y* axis of *N(t)* when *t =1*, (*ln(1) =0*); the slope *β*, is interpreted in a similar manner as a Weibull plot, If the slope *β* is greater than 1, the transmission rate is increasing, the transmission rate come more rapidly, if the slope *β* is smaller than 1, the transmission rate is decreasing, the transmission rate come more slowly, if the slope *β* is equal to 1, the process is called homogenous Poisson process, if the slope *β* is not equal *1*, the process is called Non Homogenous Poisson Process (NHPP).

Weibull distribution is invented by Dr. Waloddi Weibull in 1937, it is widely used by engineering reliability field for the failure data analysis. The slope of the Weibull plot *β* indicates which class of failures is present. CA model is also called as “Weibull Power Process” (WPP). The interpretation of the slope *β* is similar. However, the individual time to failure is used in Weibull, but the cumulative times is used in CA. Weibull distribution handles one failure mode at a time, but CA handles mixtures of situation. There are three methods to be used to fit the line, the regression, IEC (International Electrotechnical Commission) unbiased, and IEC MLE (Maximum Likelihood Estimation). The regression solution is not as accurate as the newer IEC unbiased and MLE methods except for very small samples. IEC method is from IEC 61164 [9].

## 4. Crow-AMSAA Data Analysis

### In China

The daily confirmed COVID19 cases and deaths in China are reported in the website in the reference [3]. The Crow-AMSAA model [equation (9) Ln to Ln] is applied for the cumulative total confirmed cases in China [Fig. 3]. The time period is from 1/22/2020 to 4/9/2020. It is obvious the piece-wise Crow-AMSAA can be used to fit the data. It is very interesting to see there are three phases for the COVID 19 infection. The first phase (1/22/2020 to 2/11/2020) is the growth stage where CA slope *β* is 1.683 >1, and the infectious rate is increasing. The CA slope *β* of the second phase (2/12/2020 to 2/19/2020) is 0.834 < 1, and the infectious rate is decreasing. The third phase (2/19/2020 to 4/9/2020) is towarding to the saturation stage where CA slope *β* is 0.092 < 0.834 (second phase slope *β*) <1. Chinese government locked down Hubei, Wuhan on 1/22/2020, the 14 days’ isolation of the individuals who had the contact with the COVID19 infected people, staying at home and social distance/wearing mask policy were implemented all over the country. From the CA slope *β* values (phase (1)1.683—phase (2) 0.834—phase (3) 0.092), the locking down, isolation, staying home and social distance/wearing masks played an important role to slow down the COVID19 spreading in China.

**Fig 1.**
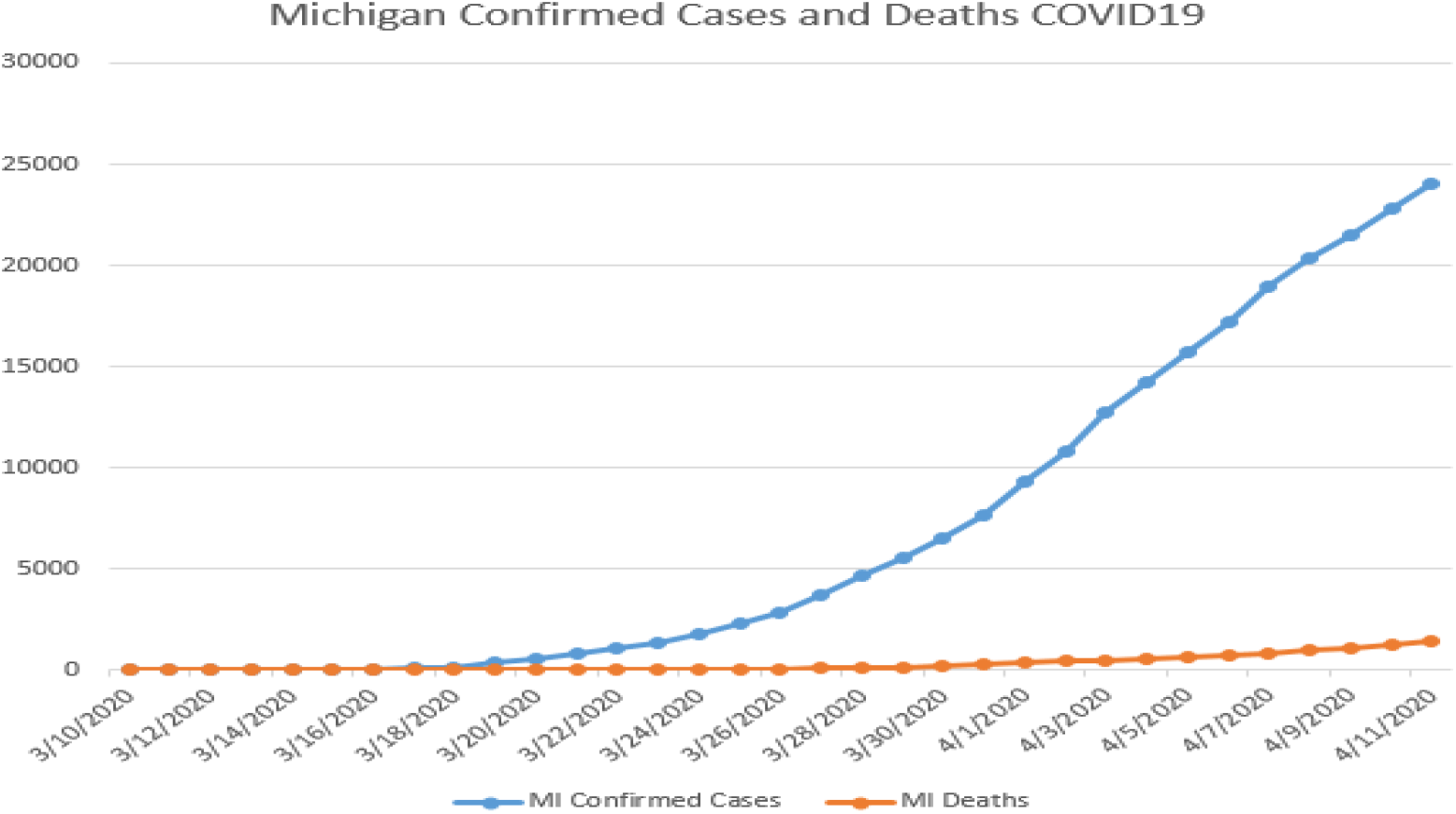
Michigan Daily Confirmed Cases and Deaths.

**Fig. 2:**
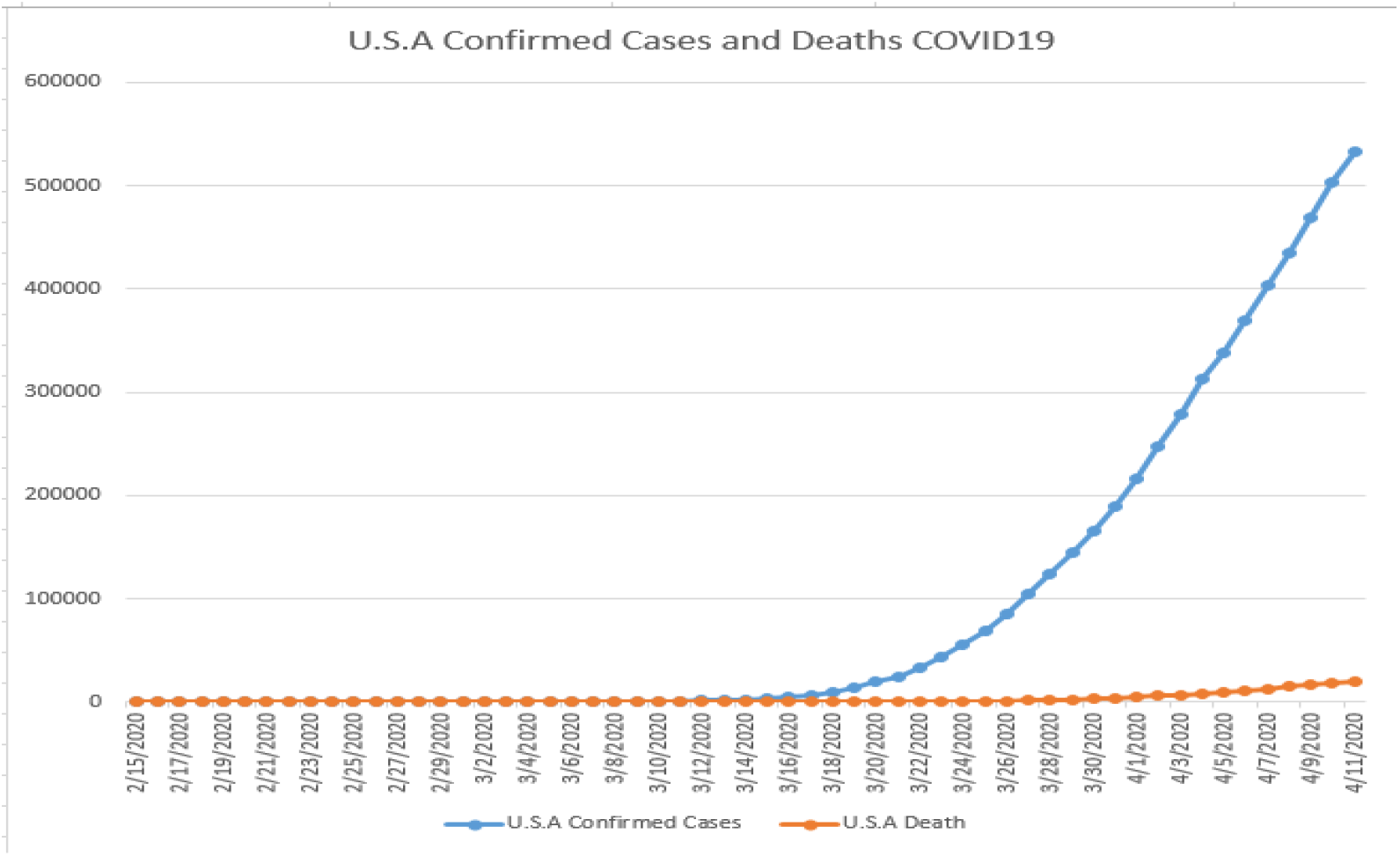
The U.S.A. Daily Confirmed Cases and Deaths.

**Fig 3.**
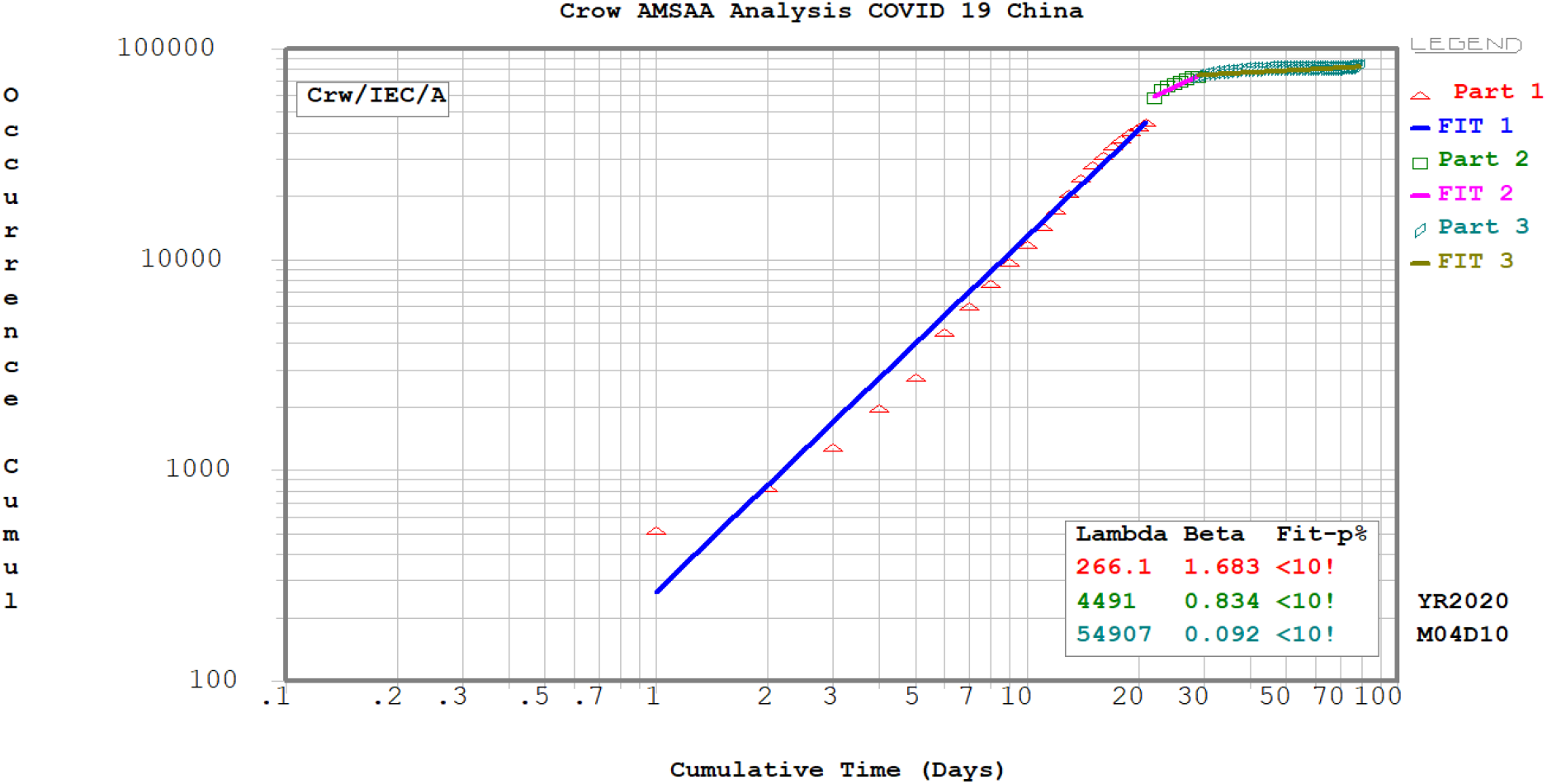
The piece wise Crow-AMSAA analysis for COVID 19 – China.

The daily death rate for COVID19 in China is plotted in Fig. 4 by using CA method. The death rate also shows the three phases. The first phase (1/22/2020 to 2/23/2020) is the death rate increasing phase where CA slope *β* is 1.829 >1. The second phase (2/24/2020 to 3/5/2020) and the third phase (3/6/2020 to 4/9/2020) are the death rate decreasing phases, the CA slopes are 0.514 and 0.141 respectively.

**Fig 4.**
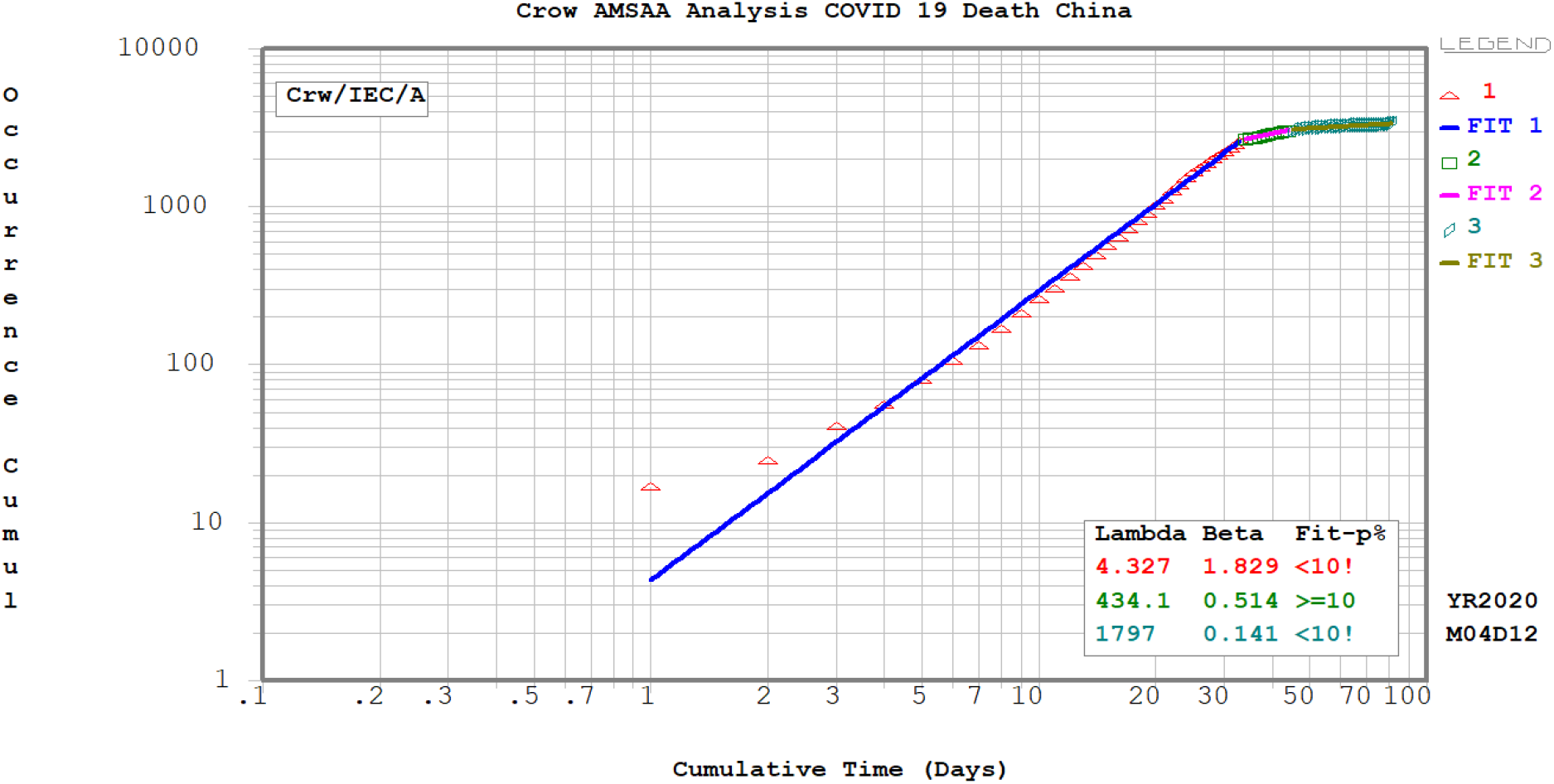
The piece wise Crow-AMSAA analysis for COVID 19 Deaths – China.

### In Michigan

The Crow-AMSAA method [equation (9) Ln to Ln] is also applied for Michigan cumulative total confirmed cases [Fig. 5]. The time period is from 3/10/2020 to 4/10/2020. So far, there are two piece wise Crow-AMSAA lines can be applied for Michigan cases. From 3/10/2020 to 3/31/2020, the CA slope *β* is 3.901 >1, and the infectious rate is increasing dramatically. From 4/1/2020 to 4/10/2020, the CA slope *β* is 2.467 >1, and the infectious rate is still increasing, though the slope *β* is slight smaller than the first phase. Since 3/24/2020, Michigan Governor issued the staying home order, the order is absolutely helping the state of Michigan to slow down the spreading of the disease, because the CA slope *β* is still greater than 1, so the infectious rate is still increasing.

**Fig 5.**
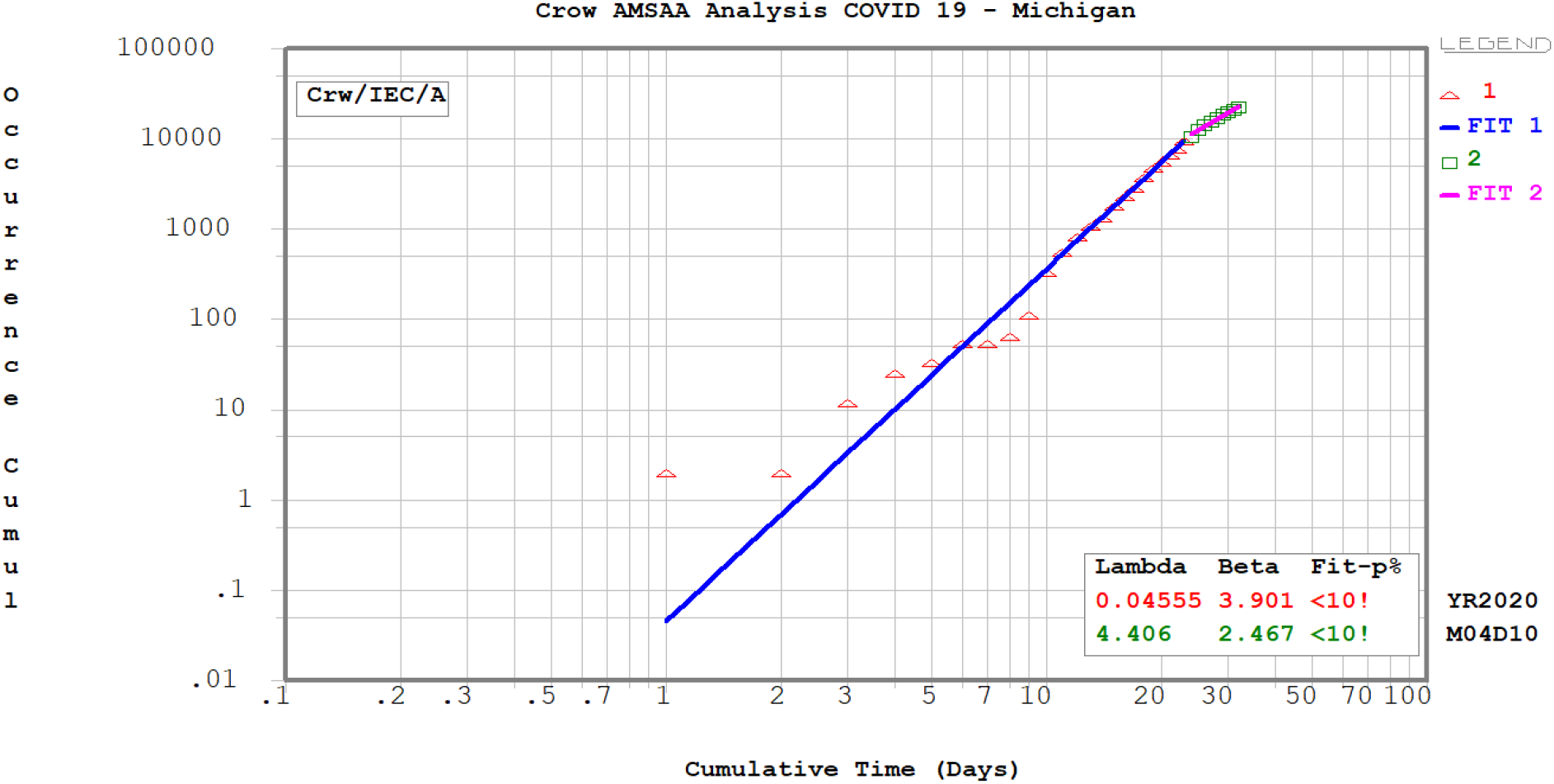
The piece wise Crow-AMSAA analysis for COVID 19 –Michigan.

The daily death rate for COVID19 in Michigan is plotted in Fig. 6 by using CA method. So far, the death rate shows the two piece of CA plots. The first piece (3/18/2020 to 3/30/2020) is the death rate increasing phase where CA slope *β* is 5.588 >1. The death rate in the second piece (3/31/2020 to 4/10/2020) is slowing down comparing to the first phase but it is still the increasing phase where the CA slopes *β* is 3.998.

**Fig 6.**
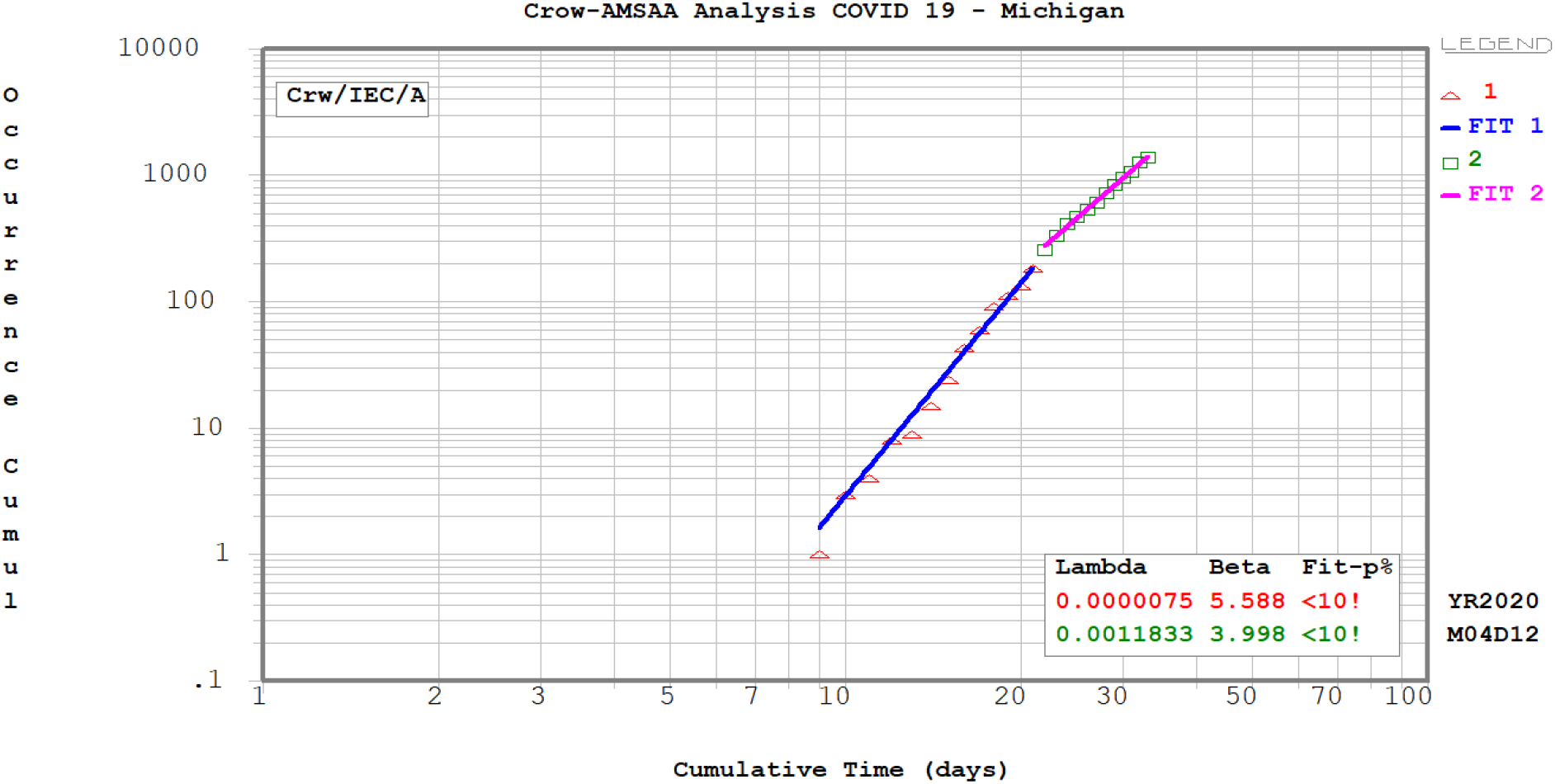
The piece wise Crow-AMSAA analysis for COVID 19 Deaths –Michigan.

### In U.S.A

The same study was conducted for U.S.A total confirmed cases [Fig.7]. From the piece-wise Crow-AMSAA plots, there are three phases so far for the U.S.A infectious cases. The first phase (2/15/2020 to 3/12/2020), the CA slope *β* is 5.138 > 1, and the infectious rate is increasing. The CA slope *β* of the second phase (3/13/202 to 3/23/2020) is 10.48 > 1, the infectious rate is increasing dramatically. The CA slope *β* of the third phase (3/24/2020 to 4/10/2020) is 5.259 >1 where the infectious rate is still increasing. Most of states in U.S.A have issued the staying at home order and social distance requirement, this will help to slow down the transmission speed of the disease.

**Fig 7.**
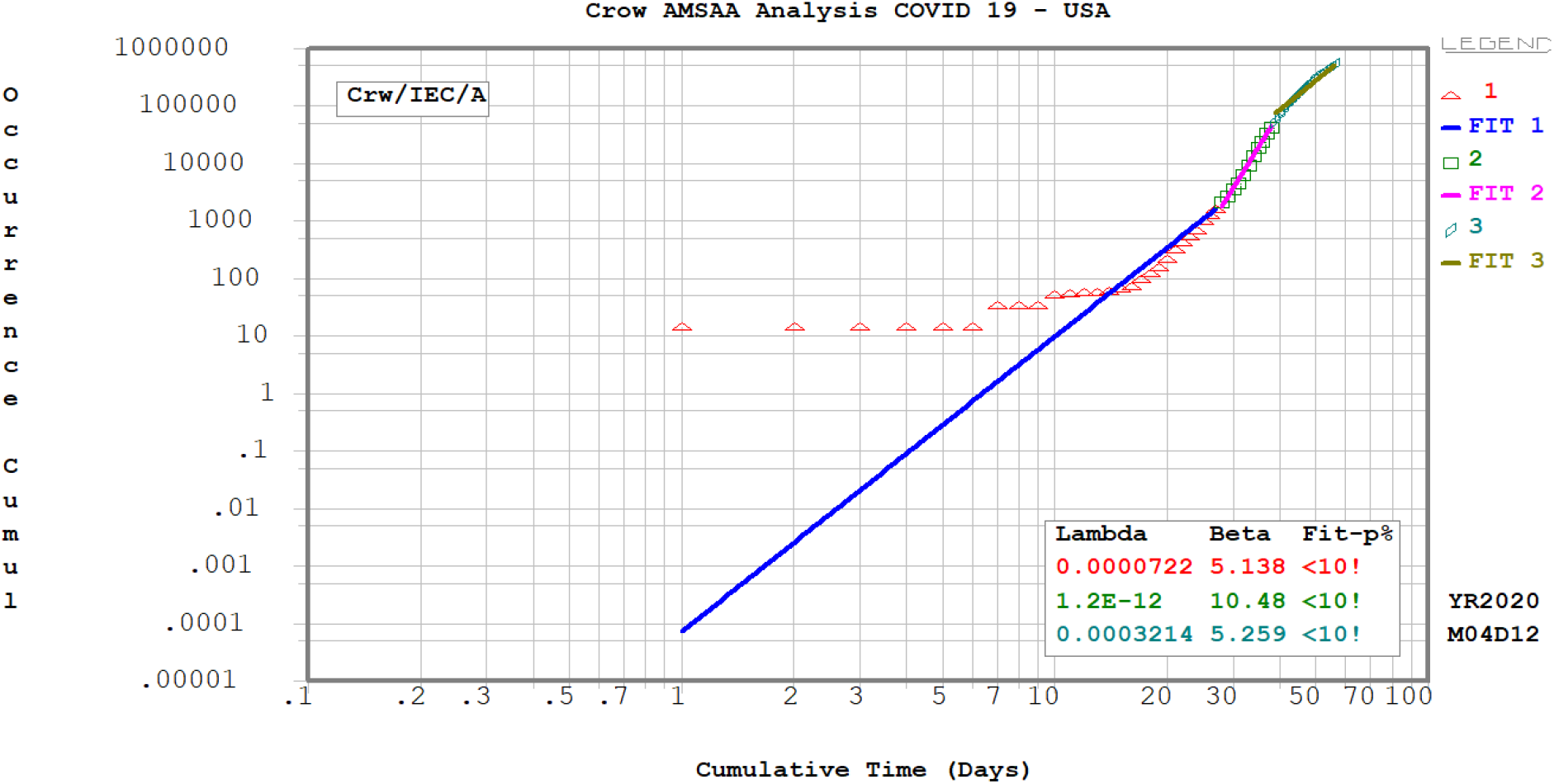
The piece wise Crow-AMSAA analysis for COVID 19 –U.S.A.

The daily death rate for COVID 19 for U.S.A is also plotted in Fig. 8. So far there are three phases identified in the plot. The CA slope *β* is 4.977 for phase I (2/19/2020 to 3/16/2020). The CA slope *β* is 10.54 for phase II (3/17/2020 to 3/27/2020) where the death rate increasing dramatically. The CA slope *β* is 7.267 for phase III (3/27/2020 to 4/11/2020) where the death rate is slowing down but it is still increasing.

**Fig 8.**
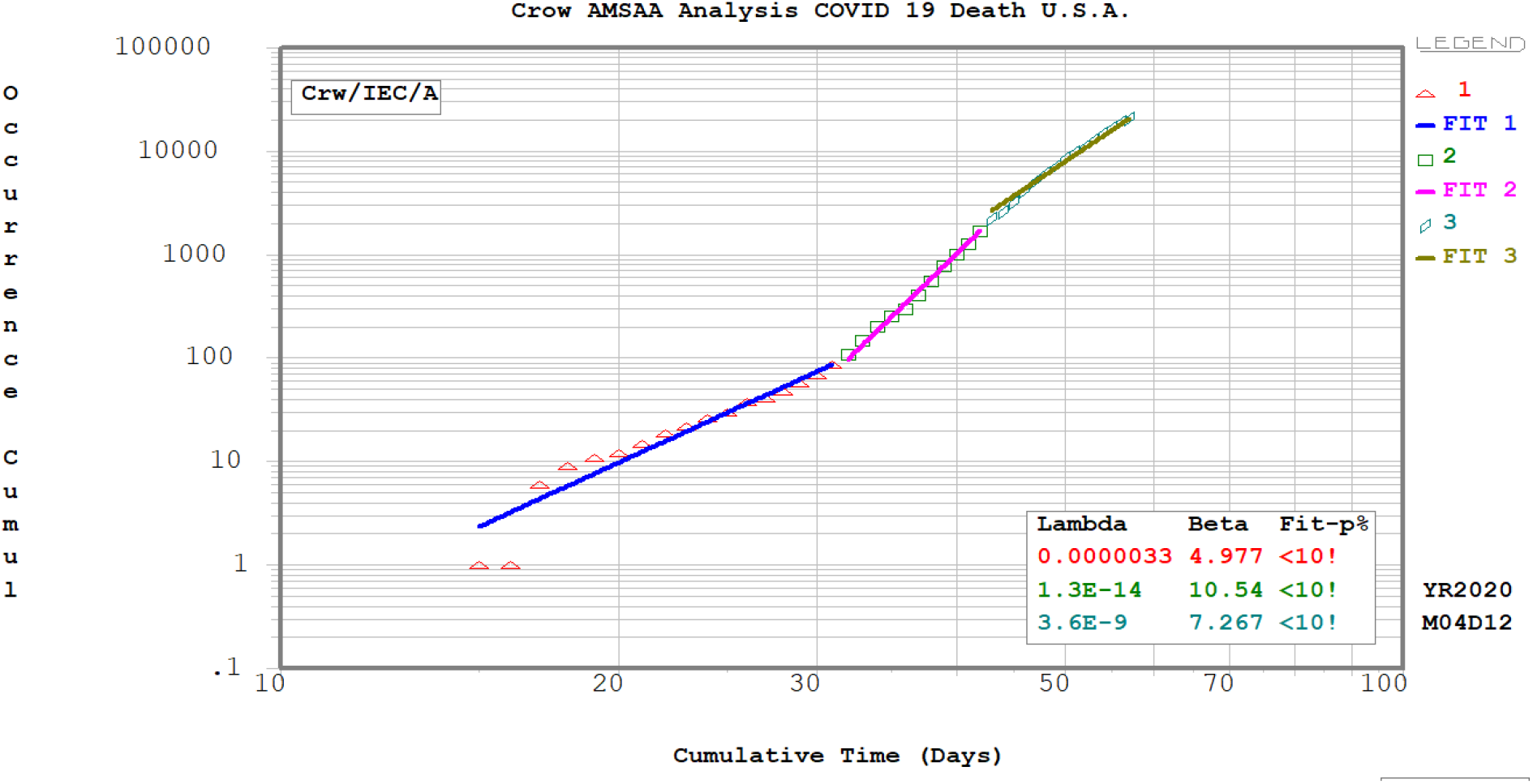
The piece wise Crow-AMSAA analysis for COVID 19 Deaths –U.S.A.

### New York City and other countries

The piece wise Crow-AMSAA analysis has been conducted for the daily confirmed cases and deaths for New York City, Spain, Italy, France, Germany, UK and South Korea [Fig. 9 to Fig. 21]. The slope *β*s are summarized in the Table 1.

**Table 1.** Summary of Crow-AMSAA slope β for different places at different phases.

| | | $\beta > 1$ rate increasing, $\beta < 1$ rate decreasing, current $\beta < \text{previous } \beta$, the rate slow down, Current $\beta > \text{previous } \beta$, the rate speed up | | |
| --- | --- | --- | --- | --- |
| | | Infectious slope $\beta$ | Death Slope $\beta$ | Current Status |
| **U.S.A** | Phase 1 | 5.138 | 4.977 | Infectious rate slow down. Death rate slow down |
|  | Phase 2 | 10.48 | 10.54 |  |
|  | Phase3 | 5.259 | 7.267 |  |
| **Spain** | Phase 1 | 7.917 | 9.284 | Infectious rate slow down. Death rate slow down |
|  | Phase 2 | 6.19 | 6.293 |  |
|  | Phase3 | 2.581 | 2.683 |  |
| **Italy** | Phase 1 | 4.18 | 5.462 | Infectious rate slow down. Death rate slow down |
|  | Phase 2 | 1.969 | 3.762 |  |
|  | Phase 3 | N/A | 1.934 |  |
| **France** | Phase 1 | 5.512 | 7.064 | Infectious rate slow down. Death rate slow down |
|  | Phase 2 | 4.796 | 5.136 |  |
|  | Phase 3 | 2.968 | N/A |  |
| **Germany** | Phase 1 | 6.909 | 8.208 | Infectious rate slow down. Death rate slow down |
|  | Phase 2 | 4.404 | 5.082 |  |
|  | Phase 3 | 2.096 | N/A |  |
| **UK** | Phase 1 | 6.567 | 8.556 | Infectious rate slow down. Death rate slow down |
|  | Phase 2 | 5.344 | 7.166 |  |
| **China** | Phase 1 | 1.683 | 1.829 | Infectious rate decreasing. Death rate decreasing. current both $\beta < 1$ |
|  | Phase 2 | 0.834 | 0.514 |  |
|  | Phase 3 | 0.092 | 0.141 |  |
| **S. Korea** | Phase 1 | 3.052 | 2.563 | Infectious rate decreasing, current $\beta < 1$, Death rate slow down |
|  | Phase 2 | 2.184 | 1.704 |  |
|  | Phase 3 | 0.393 | 1.517 |  |
| **Michigan** | Phase 1 | 3.901 | 5.588 | Infectious rate slow down. Death rate slow down |
|  | Phase 2 | 2.467 | 3.998 |  |
| **New York City** | Phase 1 | 2.42 | 4.535 | Infectious rate slow down. Death rate slow down |
|  | Phase 2 | 1.474 | 3.485 |  |
|  | Phase 3 | N/A | 1.682 |  |

**Fig 9.**
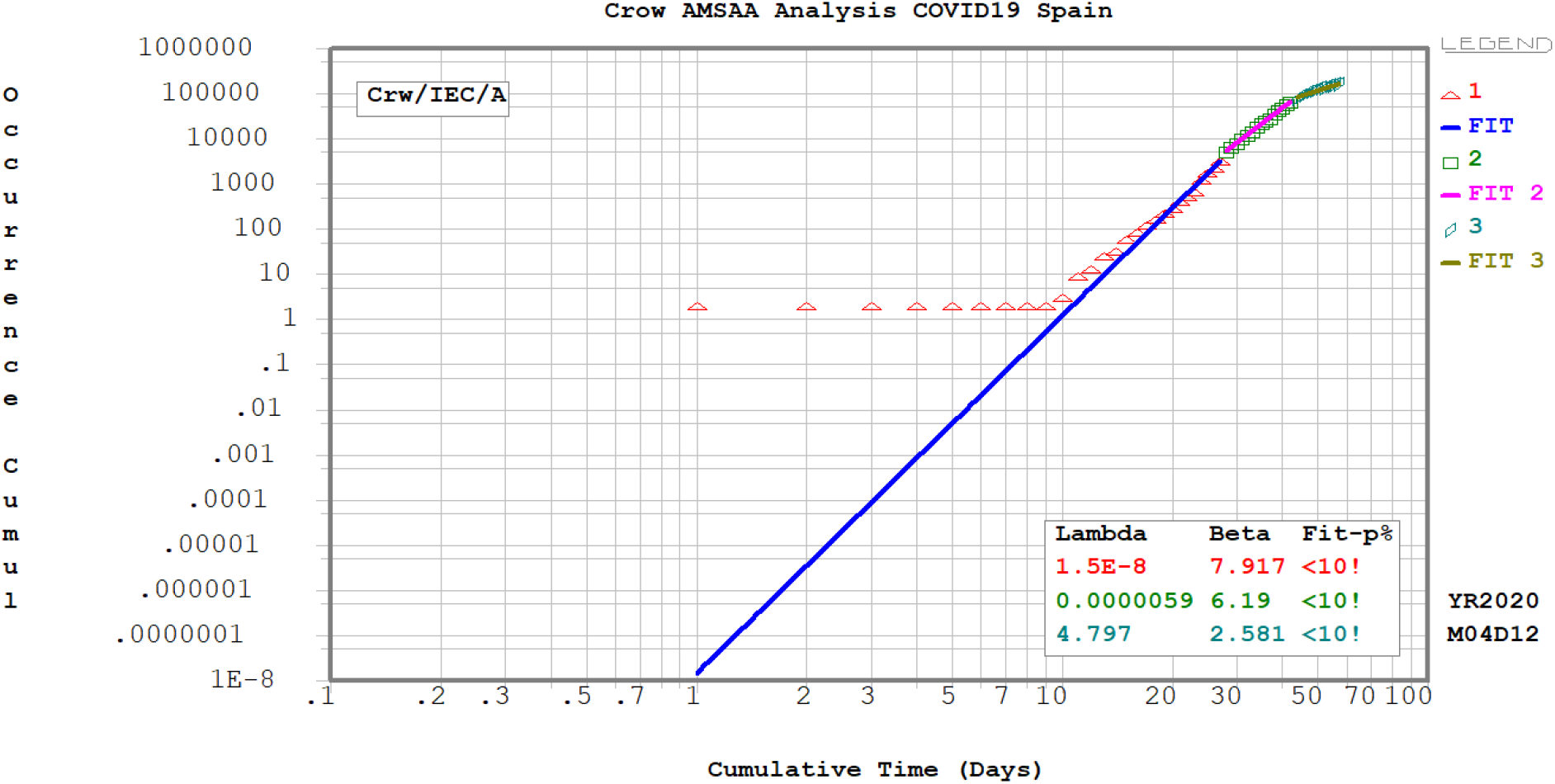
The piece wise Crow-AMSAA analysis for COVID 19 –Spain.

**Fig 10.**
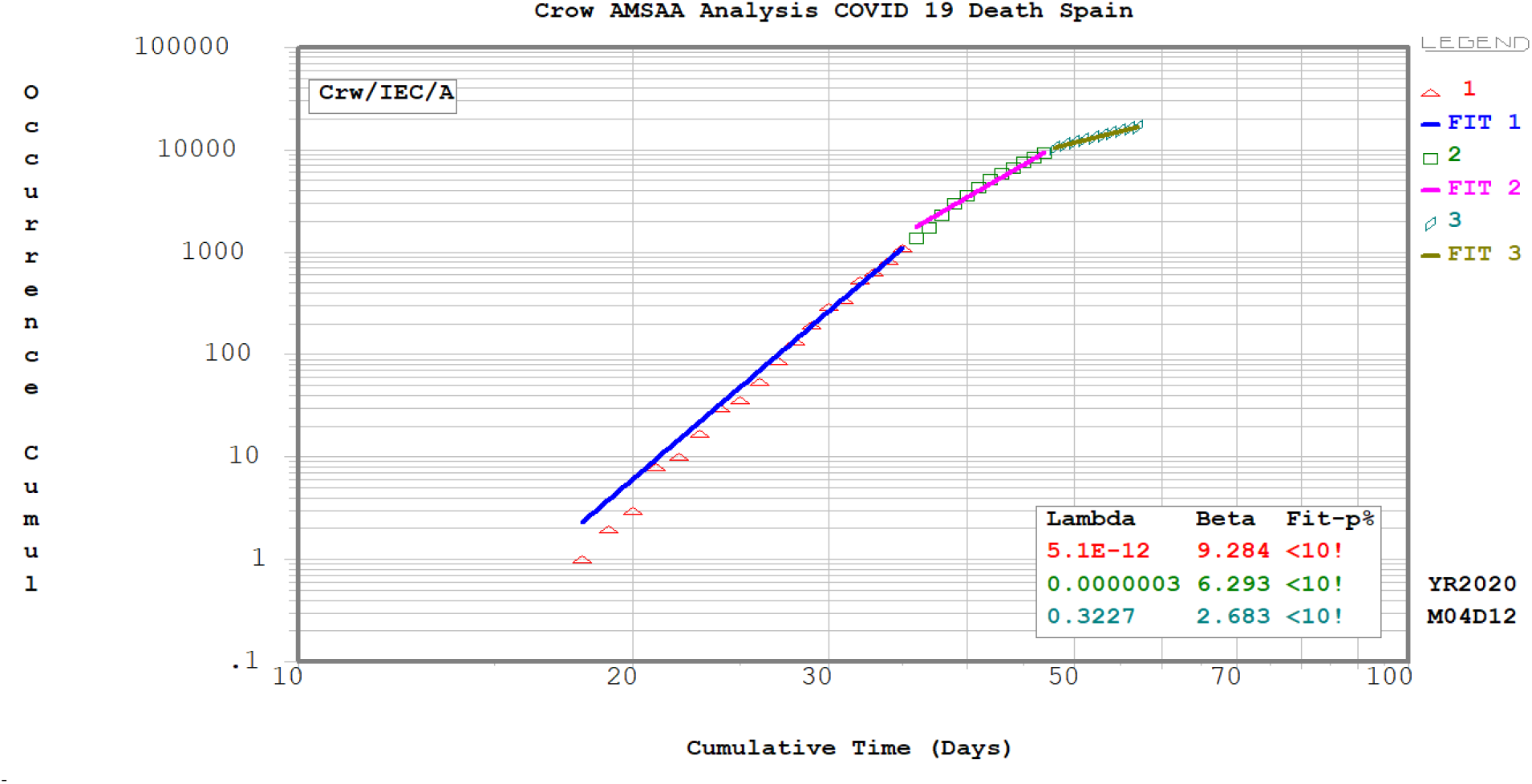
The piece wise Crow-AMSAA analysis for COVID 19 Deaths –Spain.

**Fig 11.**
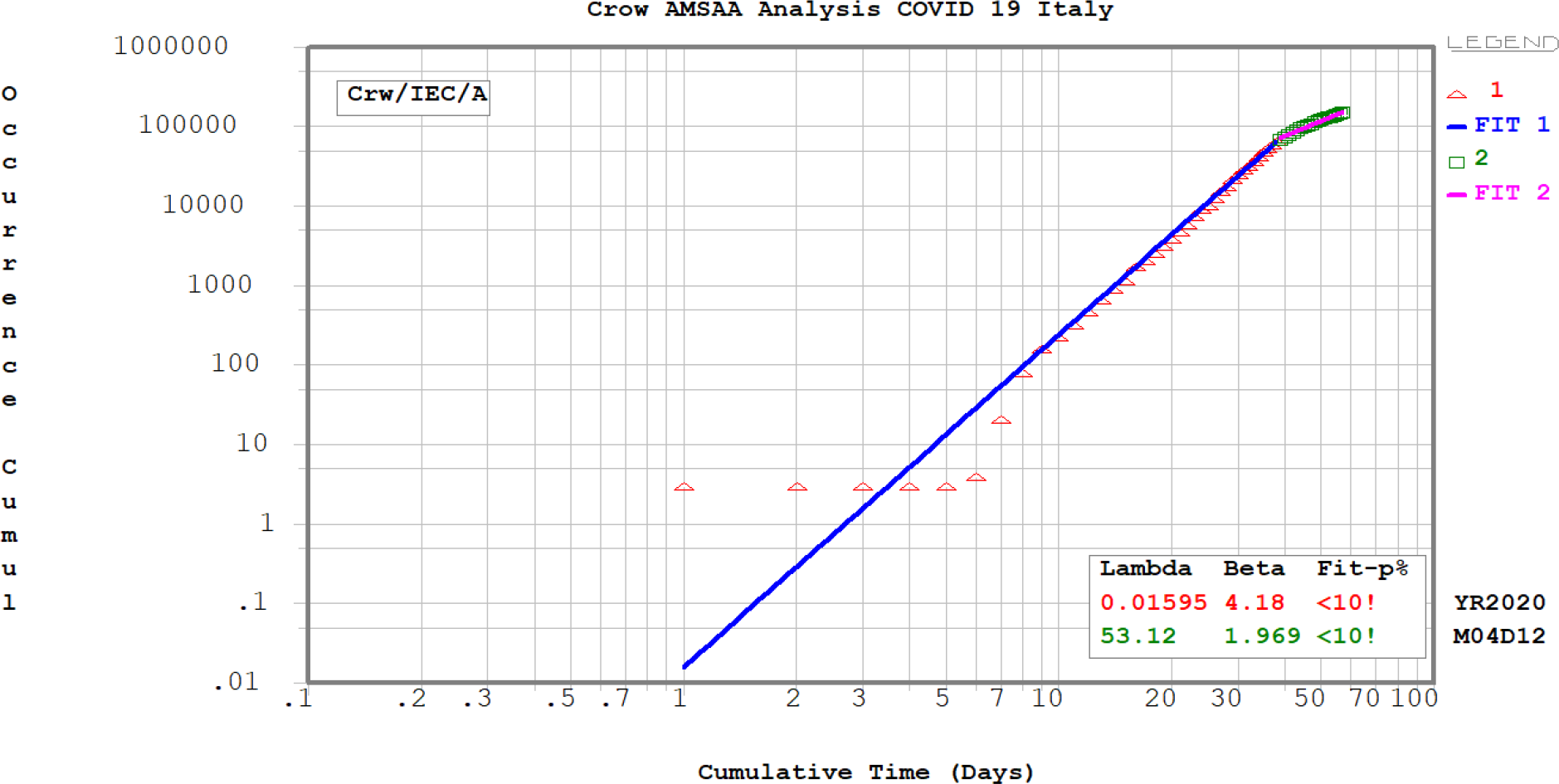
The piece wise Crow-AMSAA analysis for COVID 19 –Italy.

**Fig 12.**
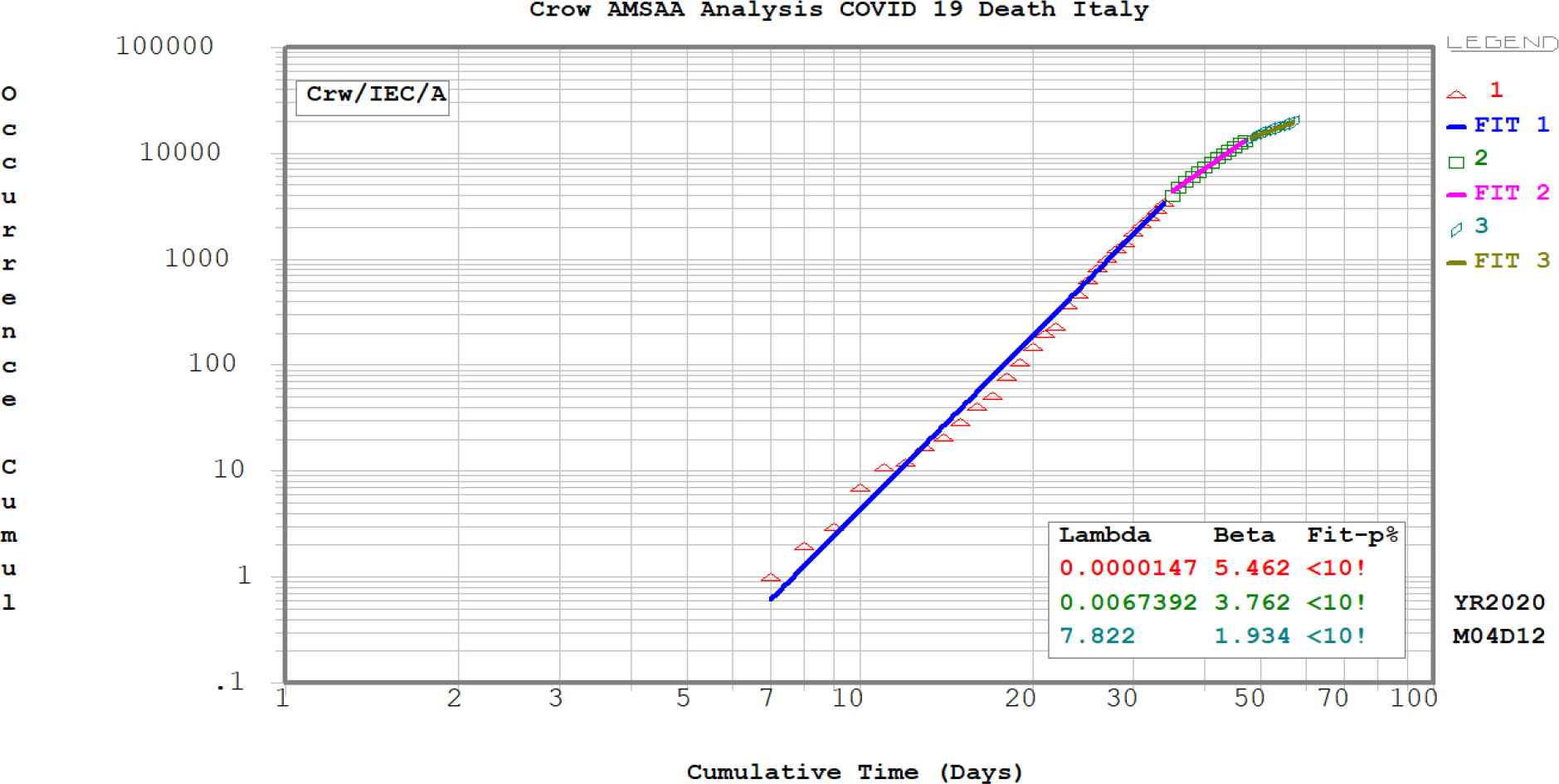
The piece wise Crow-AMSAA analysis for COVID 19 Deaths –Italy.

**Fig 13.**
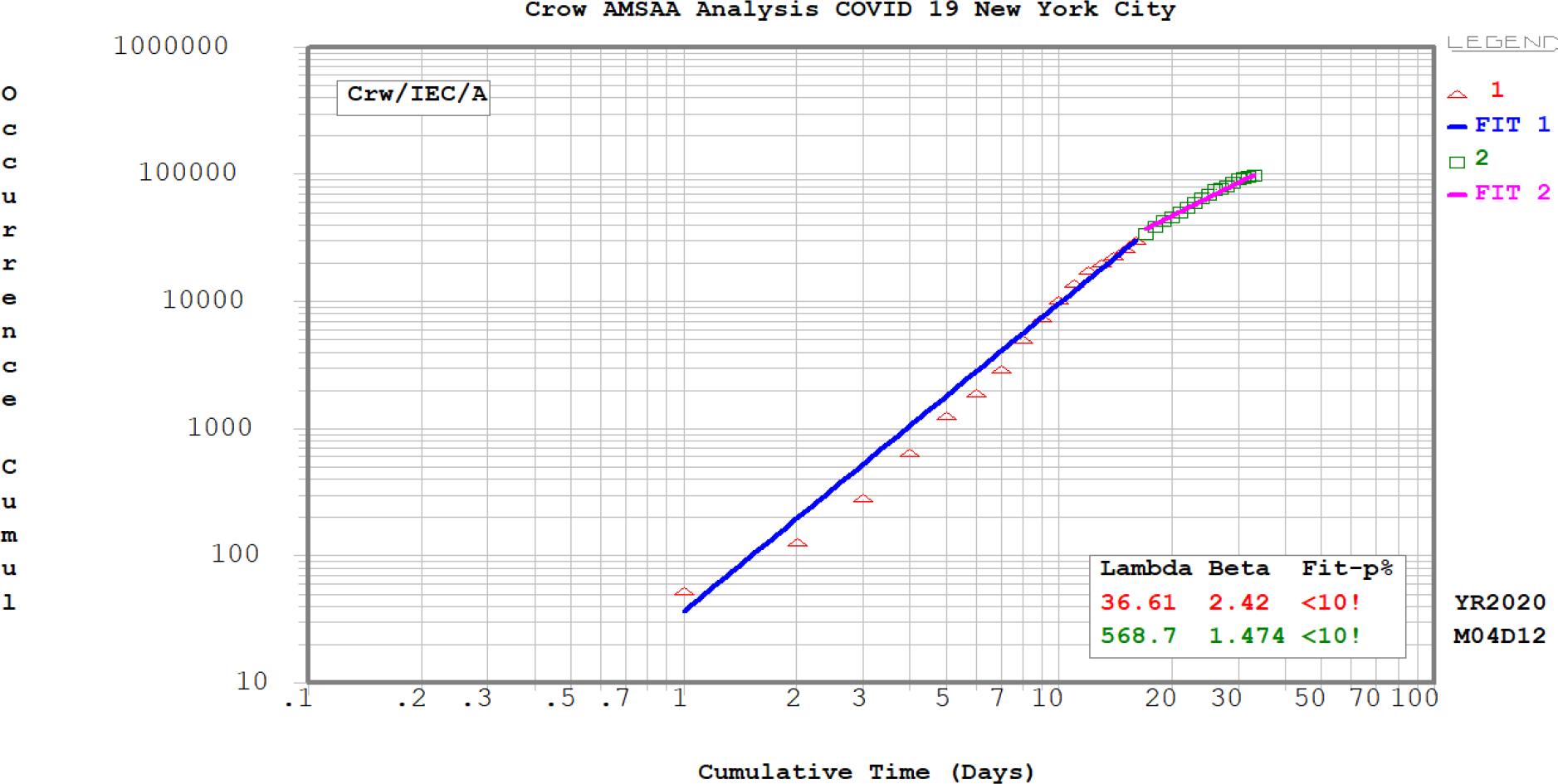
The piece wise Crow-AMSAA analysis for COVID 19 –New York City.

**Fig 14.**
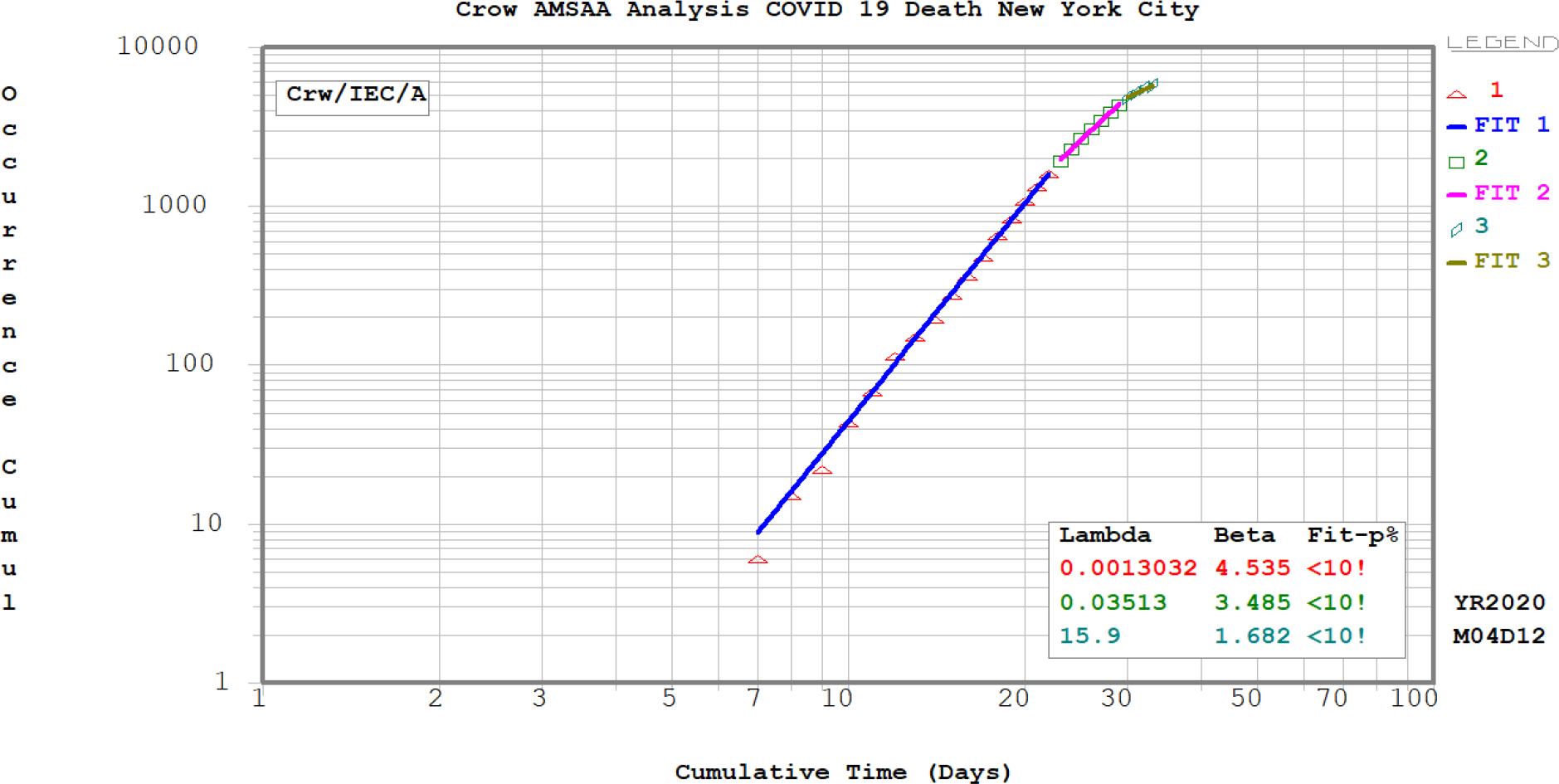
The piece wise Crow-AMSAA analysis for COVID 19 Deaths –New York City.

**Fig 15.**
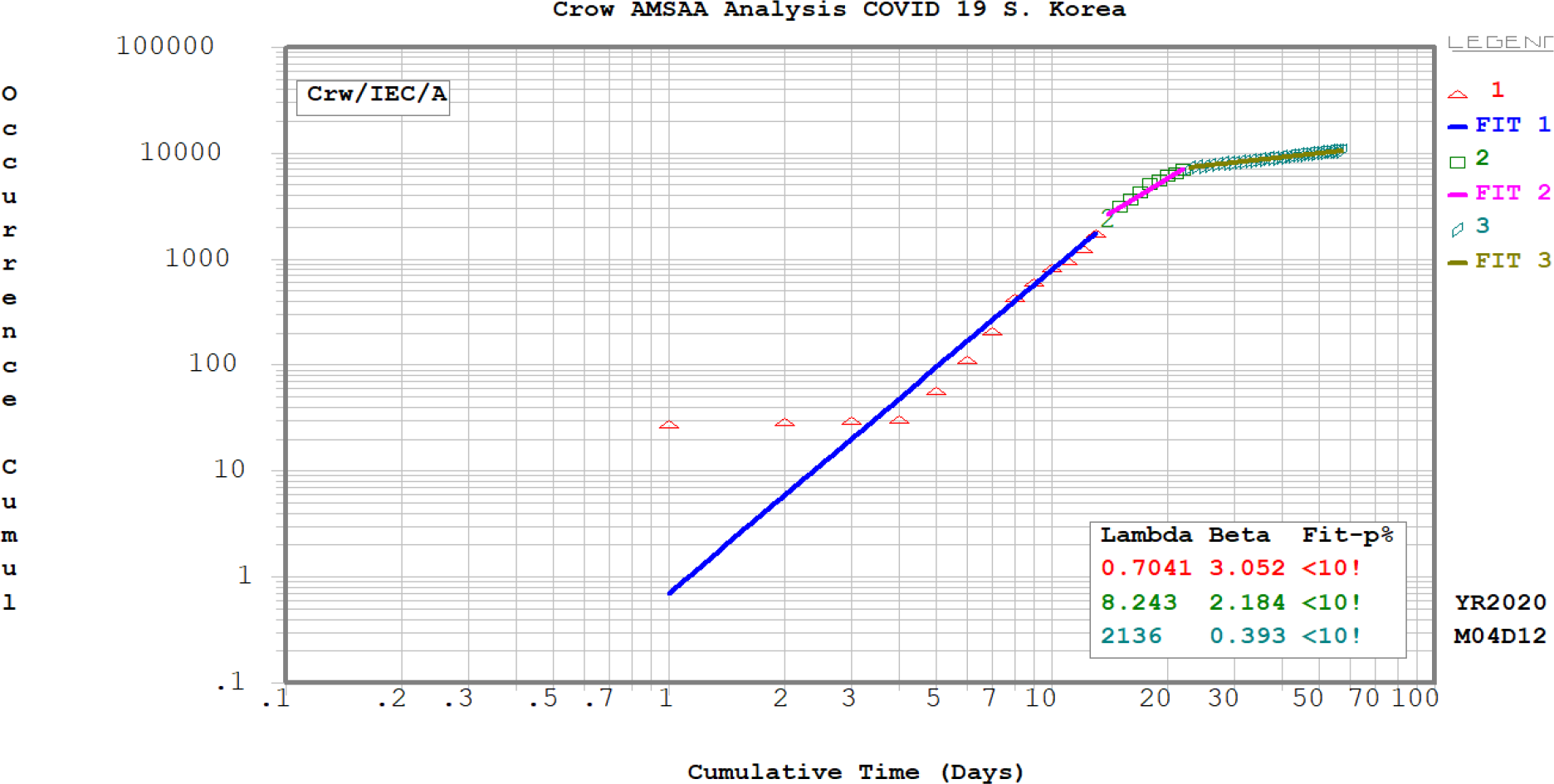
The piece wise Crow-AMSAA analysis for COVID 19 –S. Korea.

**Fig 15.**
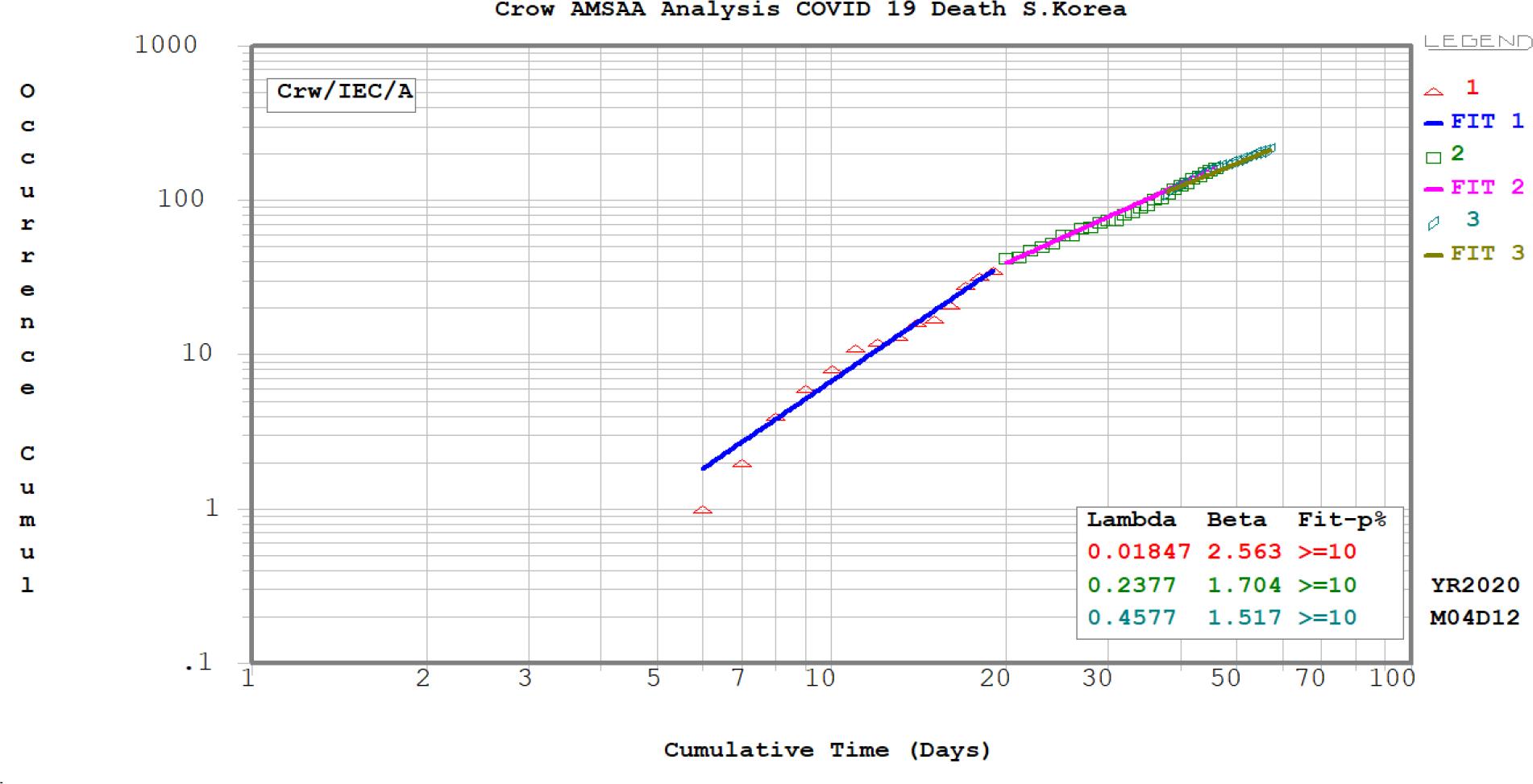
The piece wise Crow-AMSAA analysis for COVID 19 Deaths –S. Korea.

**Fig 16.**
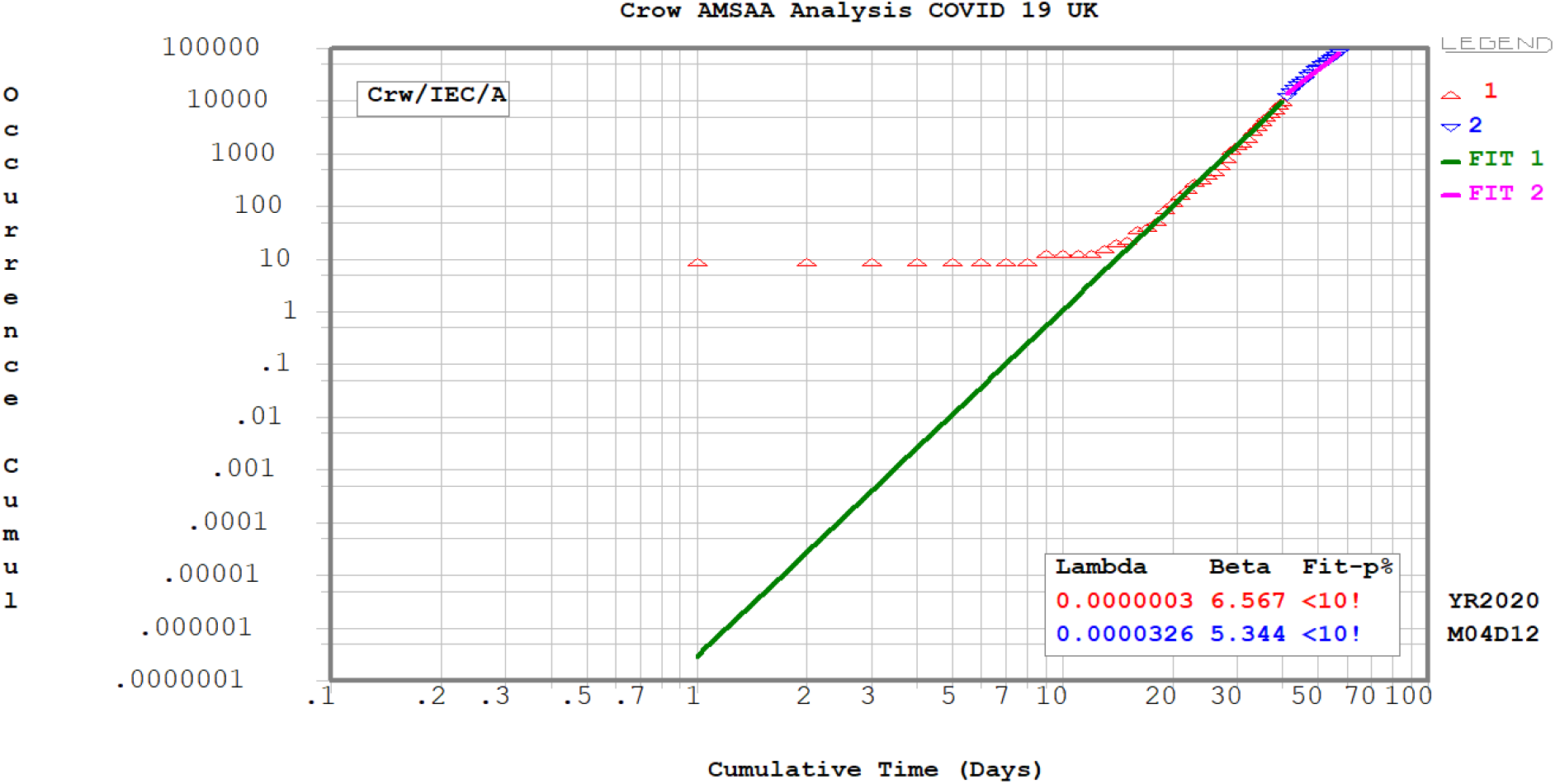
The piece wise Crow-AMSAA analysis for COVID 19 –UK.

**Fig 17.**
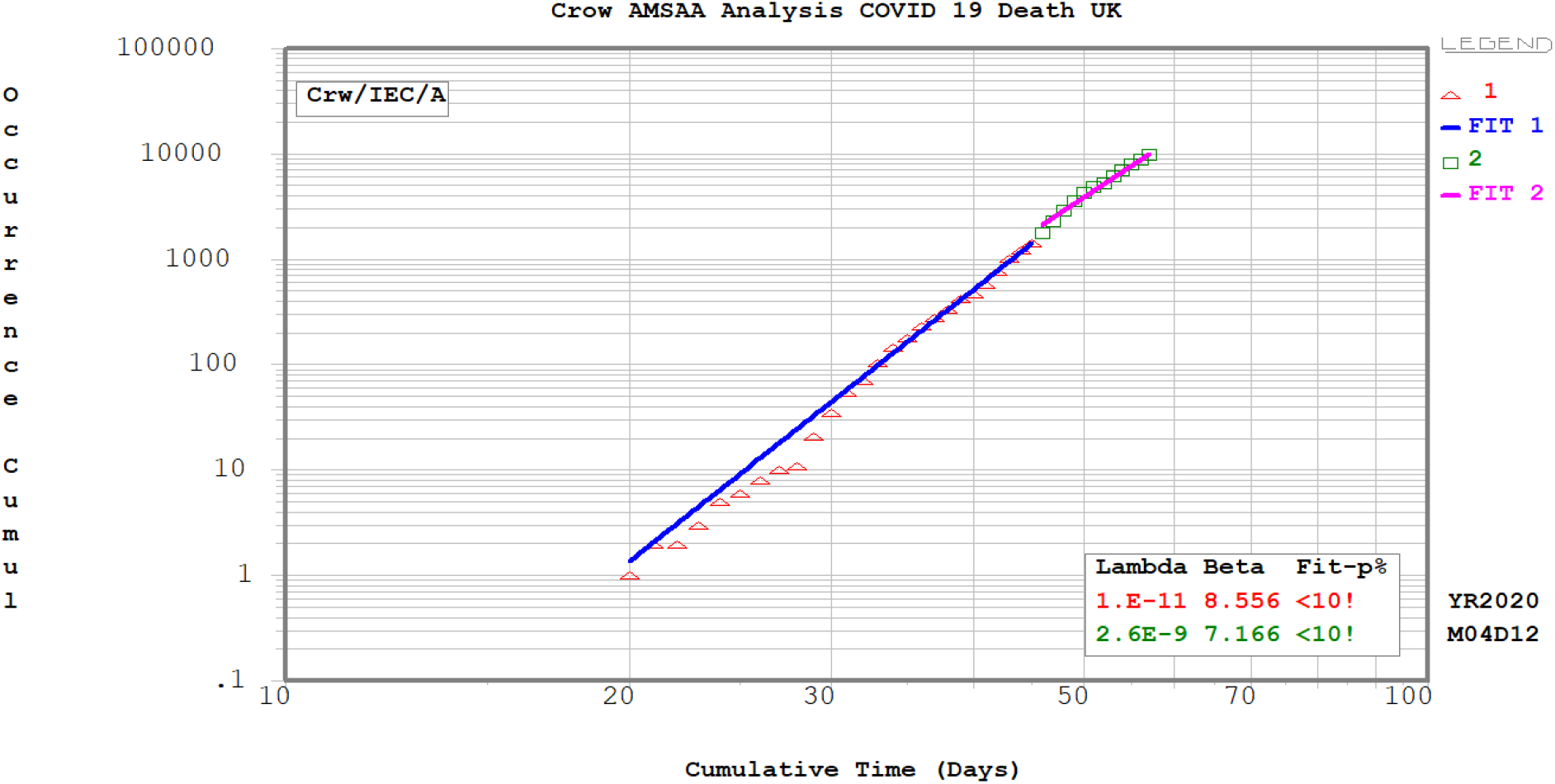
The piece wise Crow-AMSAA analysis for COVID 19 Deaths–UK.

**Fig 18.**
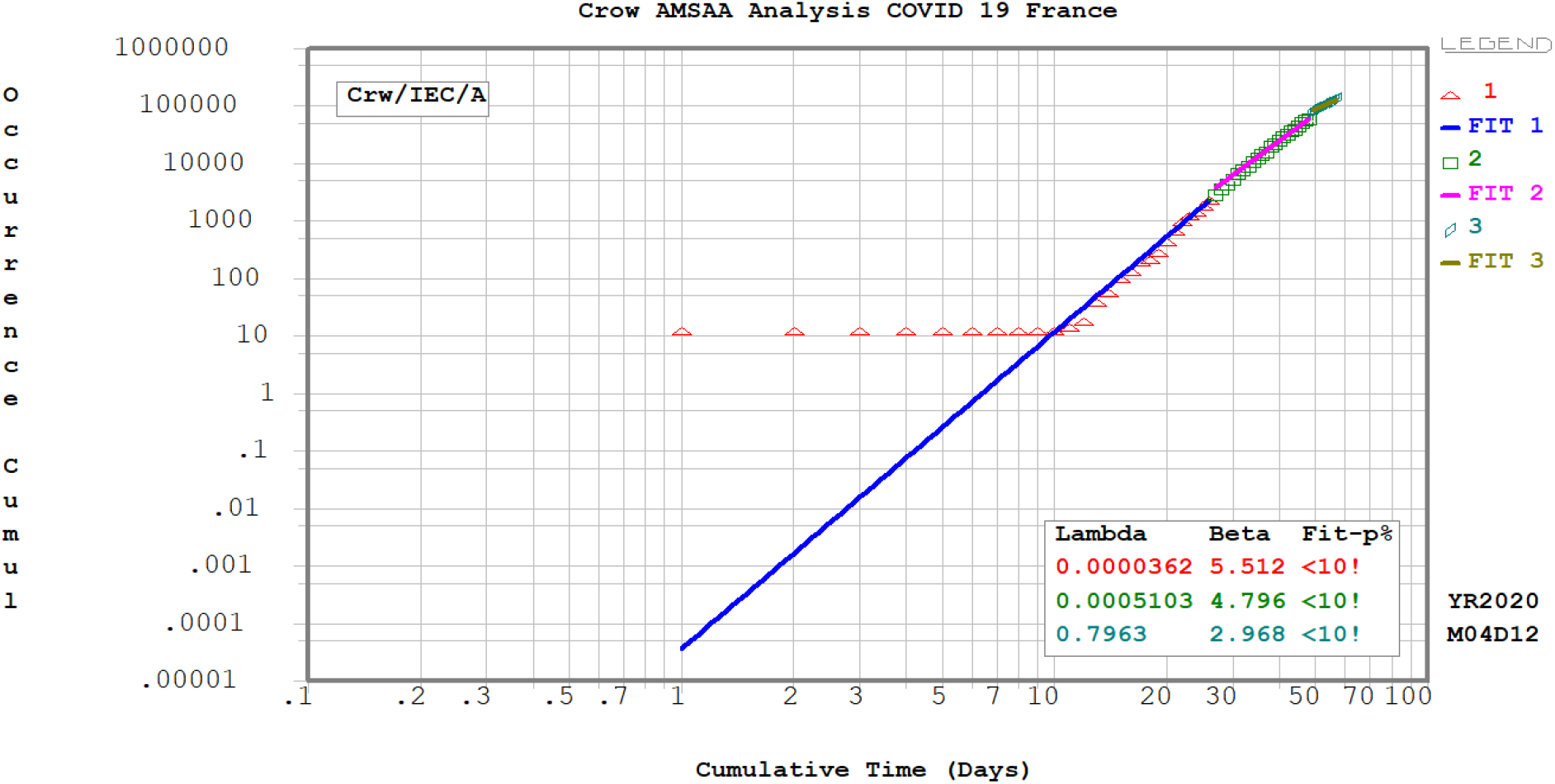
The piece wise Crow-AMSAA analysis for COVID 19 –France.

**Fig 19.**
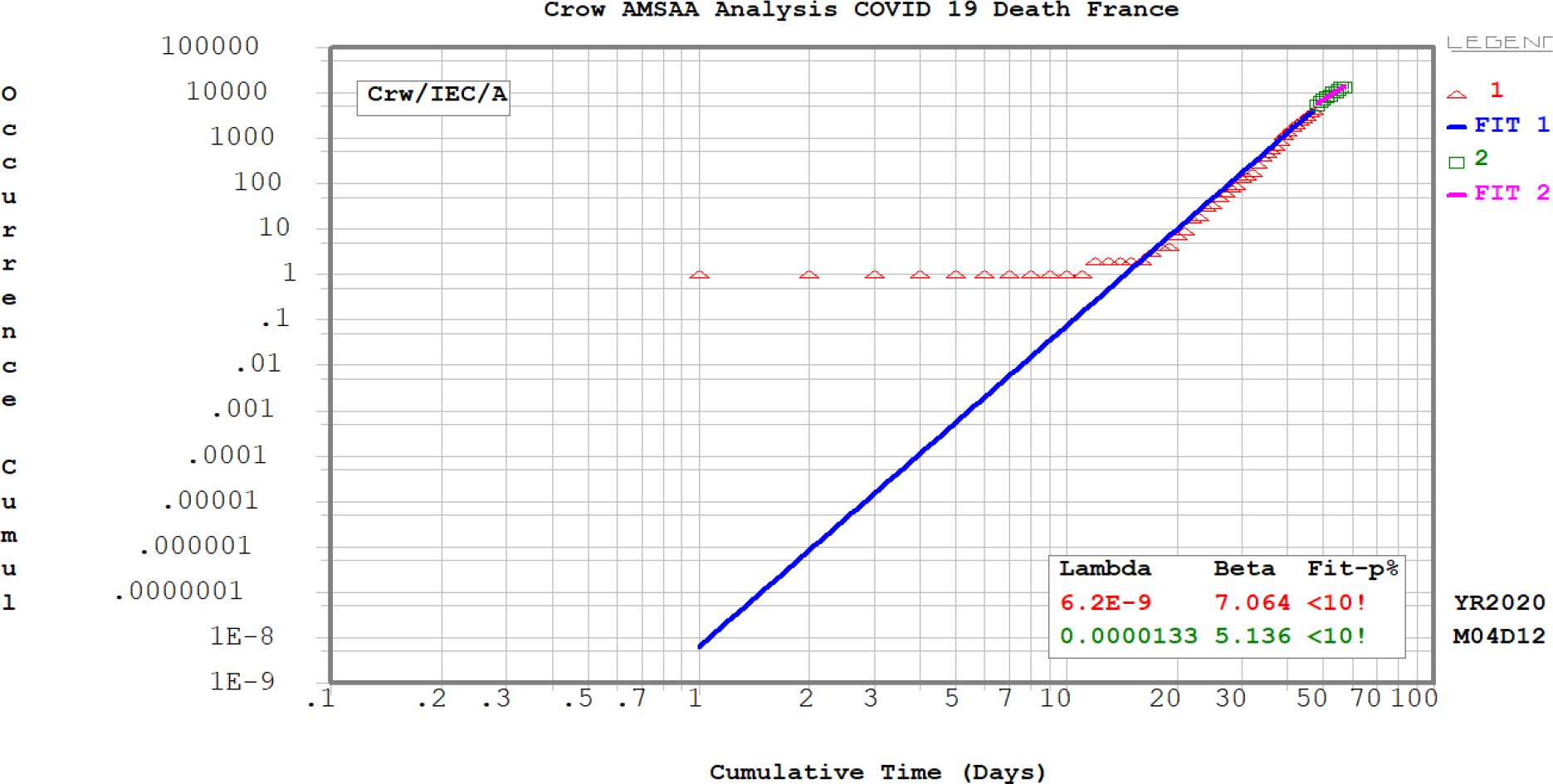
The piece wise Crow-AMSAA analysis for COVID 19 Deaths –France.

**Fig 20.**
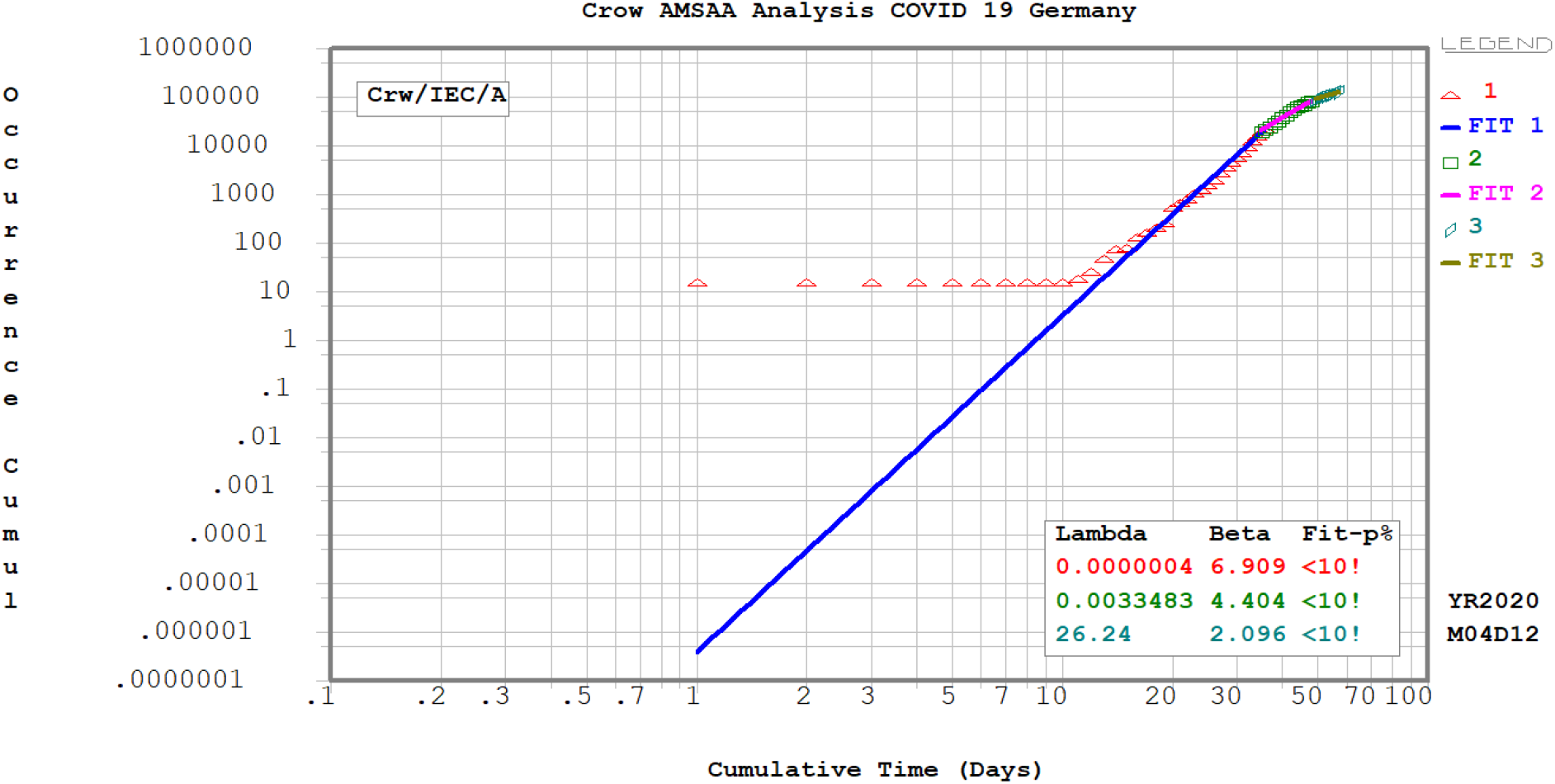
The piece wise Crow-AMSAA analysis for COVID 19 –Germany.

**Fig 21.**
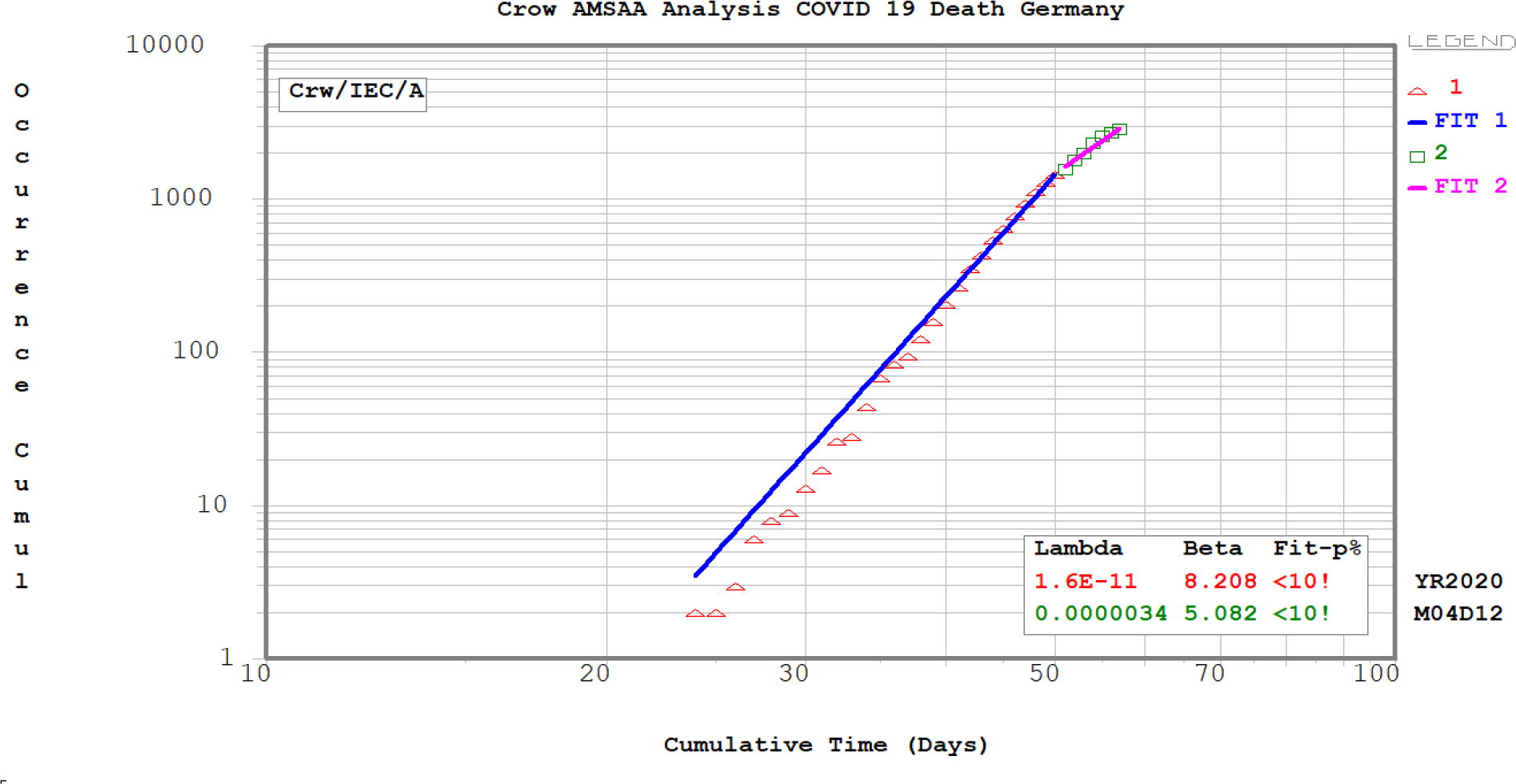
The piece wise Crow-AMSAA analysis for COVID 19 Deaths –Germany.

The decreasing/increasing of the infectious rate and death rate can be figured out per CA slope *β* values.

## 5. Discussion

From the Crow-AMSAA analysis above, at the beginning of the COVID 19, the infectious cases does not follow the Crow-AMSAA line, but when the outbreak starts, the confirmed cases does follow the CA line, the slope *β* value indicates the speed the transmission rate or death rate. The piece wise Crow-AMSAA fitting must be used in the different phases of spreading. That means the speed of the transmission rate could change according to the government interference and social distance order or other factors. Comparing the piece wise CA *β* slopes (*β:* 1.683-- 0.834-- 0.092) in China and in U.S.A (*β:*5.138*--*10.48--5.259), the speed of transmission rate in U.S.A is much higher than the infectious rate in China. From the piece wise CA plots and summary table of the CA slope *β*s, the COVID19 spreading has the different behavior at different places and countries where the government implemented the different policy to slow down the spreading.

Ranjan [5], Canabarro, etc. [6] and Liu, etc[7] are all using the traditional epidemiological model to predict the spreading the COVID19. The author is using a novel method – Crow-AMSAA which is borrowed from engineering reliability world. The Crow-AMSAA model is different from the traditional epidemiological model. The Crow-AMSAA model is the Non-Homogeneous Poisson Process (NHPP), which is for more complex problem, and NHPP models such as those for outbreaks in social networks are often believed to provide better predictions of the benefits of various mitigation strategies such as isolation, locking down and social distance [10] [11]. The piece wise Crow-AMSAA plots are used to model the expected cumulative number of infected numbers over time, and Ln-Ln plot is to simplify the curve, and slope *β* is calculated to indicate that the infectious rate is increasing or decreasing. The traditional epidemiological models is very difficult to predict the numbers of infections when the disease spreading enters to a new different phase [5].

The limitation of this piece wise Crow-AMSAA method is that the manual separation of the data has to be applied to find out the different infection phase at different time period. The good fitting of the data is depending on the good data separation.

### Future work

More studies should be done in future for COVID19 for the distribution of demographical, zone and climate conditions by using the piece wise CA models. Also the effectiveness of the government policy which preventing the spreading of this disease need be studied more to see how that affects the CA slope *β*.

## 6 Conclusion

From the above analysis for the confirmed cases and deaths for COVID 19 in Michigan, New York city, U.S.A, China and other countries, the piece wise Crow-AMSAA method can be used to modeling the spreading of COVID19.

## Data Availability

https://www.worldometers.info/coronavirus/country/us/
https://www.clickondetroit.com/news/local/2020/03/20/michigan-covid-19-data-tracking-case-count-cases-by-county-deaths-cases-by-age-tests/

https://www.clickondetroit.com/news/local/2020/03/20/michigan-covid-19-data-tracking-case-count-cases-by-county-deaths-cases-by-age-tests/

## Acknowledgments

The author appreciates the data which provided by website in reference [3] and [4]. The author also thanks the Fulton Findings company to provide the SuperSmith package.

## Supplementary Materials

